# Mapping the Evidence on Outcomes of Childhood Out-of-Home Care: A Scoping Review of Reviews

**DOI:** 10.1101/2025.05.22.25327643

**Authors:** Richmond Opoku, Natasha Judd, Katie Cresswell, Michael Parker, Michaela James, Jonathan Scourfield, Karen Hughes, Jane Noyes, Dan Bristow, Evangelos Kontopantelis, Sinead Brophy, Natasha Kennedy

## Abstract

**Background:** Children placed in out-of-home care in high-income countries face complex challenges due to exposure to adverse childhood experiences and systemic disadvantages. While research on their outcomes has grown, the evidence base remains fragmented. An overview of review-level evidence was conducted to identify patterns, gaps, and priorities for future research and practice.

**Methods:** A scoping review of reviews was conducted. Peer-reviewed review articles published between January 2013 and July 2024 were identified through searches in databases including EBSCOhost, ProQuest, Cochrane Database of Systematic Reviews, and Epistemonikos. Eligible reviews focused on childhood out-of-home care experience and reported outcomes for care-experienced individuals (assessed either in childhood or adulthood) and/or associated factors. Outcomes were categorised under the following domains: Health and Emotional Wellbeing (HEW), Physical and Legal Security (PLS), Education and Learning (EL), Living Standards and Social Wellness (LSSW), and Identity and Civic Participation (ICP). Factors were classified across multiple levels, including individual child-level, socio-relational-level, community-level, system-level, and other factors.

**Results:** A total of 77 reviews were included, spanning diverse methodologies and contexts. Research was concentrated in domains such as HEW and LSSW, with indicators such as mental and emotional health and attachment and behaviour functioning receiving substantial attention. Conversely, key gaps were identified, including the limited reporting of ICP outcomes (e.g., identity and self-respect). System-level factors, such as care quality and placement type, were most frequently reported across outcome domains and indicators. Individual child level and socio-relational-level factors were consistently highlighted, while community-level factors were largely underrepresented.

**Conclusion:** Future research should target gaps in underexplored outcome domains like ICP and indicators such as bullying, mortality, and educational readiness. Community-level factors warrant more attention as they play a significant role in supporting transitions to independence and social integration.

## Background

In most high-income countries, current laws and policies emphasise placing children who cannot remain with their parents into out-of-home care (OHC), with a preference for placements with extended family members or close family friends (kinship care) [1–3]. Some argue that kinship care arrangements are associated with better developmental outcomes compared to placements in unrelated foster care or institutional settings [4,5]. However, children in any form of OHC remain a particularly vulnerable group, with complex needs and facing numerous challenges. These challenges often stem from adverse childhood experiences (e.g., abuse and neglect) and systemic disadvantages encountered prior to, during, and sometimes after their time in care [4,6–10].

Over the past decades, increasing attention has been given to the experiences and outcomes of children in OHC, reflected in a growing body of research, including several literature reviews [e.g., 11–16]. These reviews synthesise evidence across diverse contexts, but the research landscape remains fragmented, with studies varying significantly in focus and methodological approach. With this in mind, a comprehensive map of studied outcomes and their associated factors would provide a clearer understanding of the breadth of available evidence, help identify gaps, and highlight priority areas for future research and intervention.

This study aims to systematically map existing reviews on the outcomes of OHC and the factors examined in relation to these outcomes. By collating and analysing the available evidence, this study will serve as a resource for policymakers, practitioners, and researchers. This will ensure that efforts to support children in OHC are grounded in a robust understanding of the evidence base.

## Materials and Methods

### Review design

A scoping review of reviews was determined to be the most suitable approach for capturing the breadth of this extensive research area, systematically mapping the existing evidence, and identifying gaps in the literature. The protocol for the review is available at https://doi.org/10.17605/OSF.IO/G7D5J. The review was guided by the recommendations in Schultz [17]. Reporting followed the Preferred Reporting Items for Systematic Reviews and Meta-Analyses (PRISMA) extension for scoping reviews [18].

### Search strategy

A search of academic databases was conducted to identify peer-reviewed review articles. The databases included EBSCOhost (searching MEDLINE, APA PsycArticles, APA PsycInfo, Education Research Complete, and CINAHL Ultimate), ProQuest (covering ASSIA, Criminal Justice Database, Education Database, and Social Science Database), Cochrane Database of Systematic Reviews, and Epistemonikos. The initial search strategy was developed by R.O. and underwent iterative refinement following inputs from J.N., K.H., S.B., and Patient and Public Involvement (PPI) participants. The full search strategy, details on PPI activity, and the procedures to select studies are described in a related review of reviews [19] (see S1 File).

### Eligibility criteria

A review study was included if it met the following criteria:

- Reported any outcome(s) for individuals placed in OHC during childhood (i.e., under 18 years of age), assessed either in childhood or adulthood, and/or associated factors.
- Included quantitative, qualitative, or mixed-methods studies with a documented search strategy.
- Published from January 2013 to July 2024, available in full-text and written in English.
- Focused on global or high-income country contexts. The decision to focus on high-income countries was based on the following considerations:
- These countries are more likely to have established and formalised systems of fostering, kinship, and residential care, offering a clearer basis for cross-study comparisons.
- The review aimed to inform policy and research priorities within high-income contexts.

A review study was excluded if it:

- Focused exclusively on children with learning disabilities, those in inpatient psychiatric care, young offender institutions, adoption, or specialist centres for mothers and children. These contexts often involve distinct care pathways, interventions, and outcome profiles that differ meaningfully from the broader looked-after population, and were thus beyond the scope of this review.
- Was non-empirical or exclusively focused on literature from low- and middle-income countries or postgraduate theses, books, or grey literature.

### Charting the data

Data extraction was carried out independently by three reviewers (R.O., K.C., and N.J.), while two supervising reviewers (M.J. and S.B.) verified the extracted data to minimise errors. The following information was collected from each included review: author, year of publication, title, type of review, number and type of included studies, countries covered by the included studies, review time frame, population, analytical approach, aims/objectives, findings on factors associated with care entry, review authors’ interpretations, and quality assessments of the included studies. It is common practice in studies mapping evidence on broad topics to limit data extraction to the abstract section [17,20–23]. However, data extraction for this review was extended to include the full text to minimise the risk of underestimating the number of reviews reporting relevant outcomes and factors [24].

### Collating, summarising, and reporting results

The large volume of records, combined with the broad scope of the topic and diverse outcome measures, makes synthesising and reporting specific findings impractical [17,24]. However, a descriptive analysis of study characteristics and outcome categories enabled a comprehensive mapping the literature. One reviewer (R.O.) categorised the outcomes of children in care using the domains and indicators of the Equality Measurement Framework (EMF), developed by the UK Equality and Human Rights Commission [25]. Adaptations were made to account for the EMF not being specifically designed for children in OHC. Some domains were merged, while additional indicators emerged from the data, resulting in the five outcome domains described in Table 1. Details on the development of outcome categories are available in S2 Table. Additionally, factors were categorised using the modified version of the ecological model [7] described in Table 1. The term "factor(s)" is used broadly to encompass various influences identified by the reviews, extending beyond variables with statistical associations. This approach captured diverse influences on outcomes, as some reviews synthesised qualitative findings on barriers and facilitators, while others identified associations through statistical analyses. One reviewer (N.J.) inspected categorisation of outcomes and associated factors to ensure consistency and accuracy in how findings were grouped and interpreted.

**Table 1.**
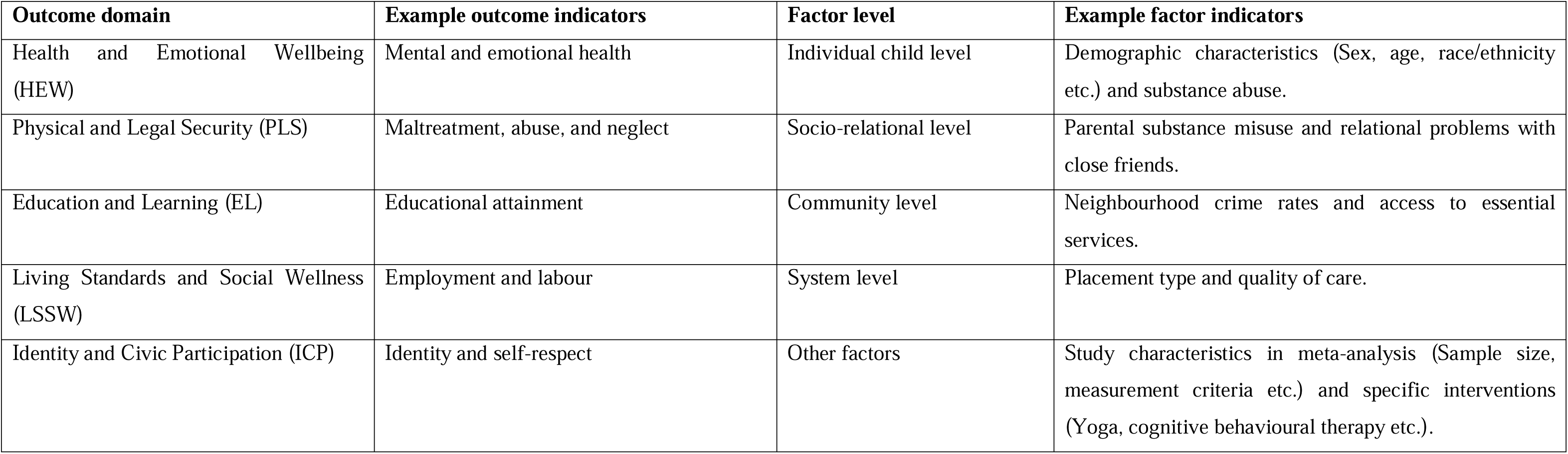
Categorisation of outcomes and factors

| **Outcome domain** | **Example outcome indicators** | **Factor level** | **Example factor indicators** |
| --- | --- | --- | --- |
| Health and Emotional Wellbeing (HEW) | Mental and emotional health | Individual child level | Demographic characteristics (Sex, age, race/ethnicity etc.) and substance abuse. |
| Physical and Legal Security (PLS) | Maltreatment, abuse, and neglect | Socio-relational level | Parental substance misuse and relational problems with close friends. |
| Education and Learning (EL) | Educational attainment | Community level | Neighbourhood crime rates and access to essential services. |
| Living Standards and Social Wellness (LSSW) | Employment and labour | System level | Placement type and quality of care. |
| Identity and Civic Participation (ICP) | Identity and self-respect | Other factors | Study characteristics in meta-analysis (Sample size, measurement criteria etc.) and specific interventions (Yoga, cognitive behavioural therapy etc.). |

## Results

### Study selection and characteristics

Figure 1 presents detailed data on the study selection process. Of the 711 references initially imported, 656 were screened by title and abstract, leading to 143 full-text assessments. Finally, 77 studies met the eligibility criteria for inclusion in the review. Among these studies, 38 (49.4%) were systematic reviews [11,12,26–61], 10 (13.0%) were meta-analyses [16,62–70], seven (9.1%) were systematic reviews with meta-analyses [5,71–76], six (7.8%) were scoping reviews [13,14,77–80], and 16 (20.8%) were other review types [15,81–95]. The reviews included primary studies from at least 48 countries, with most conducted in the USA, followed by the UK, Canada, Australia, and Sweden. The primary studies spanned the years 1972 to 2022. Over 60% of the reviews assessed either the risk of bias or the quality of the included primary studies. The full characteristics of the included reviews are presented in S3 Table.

**Figure 1.**
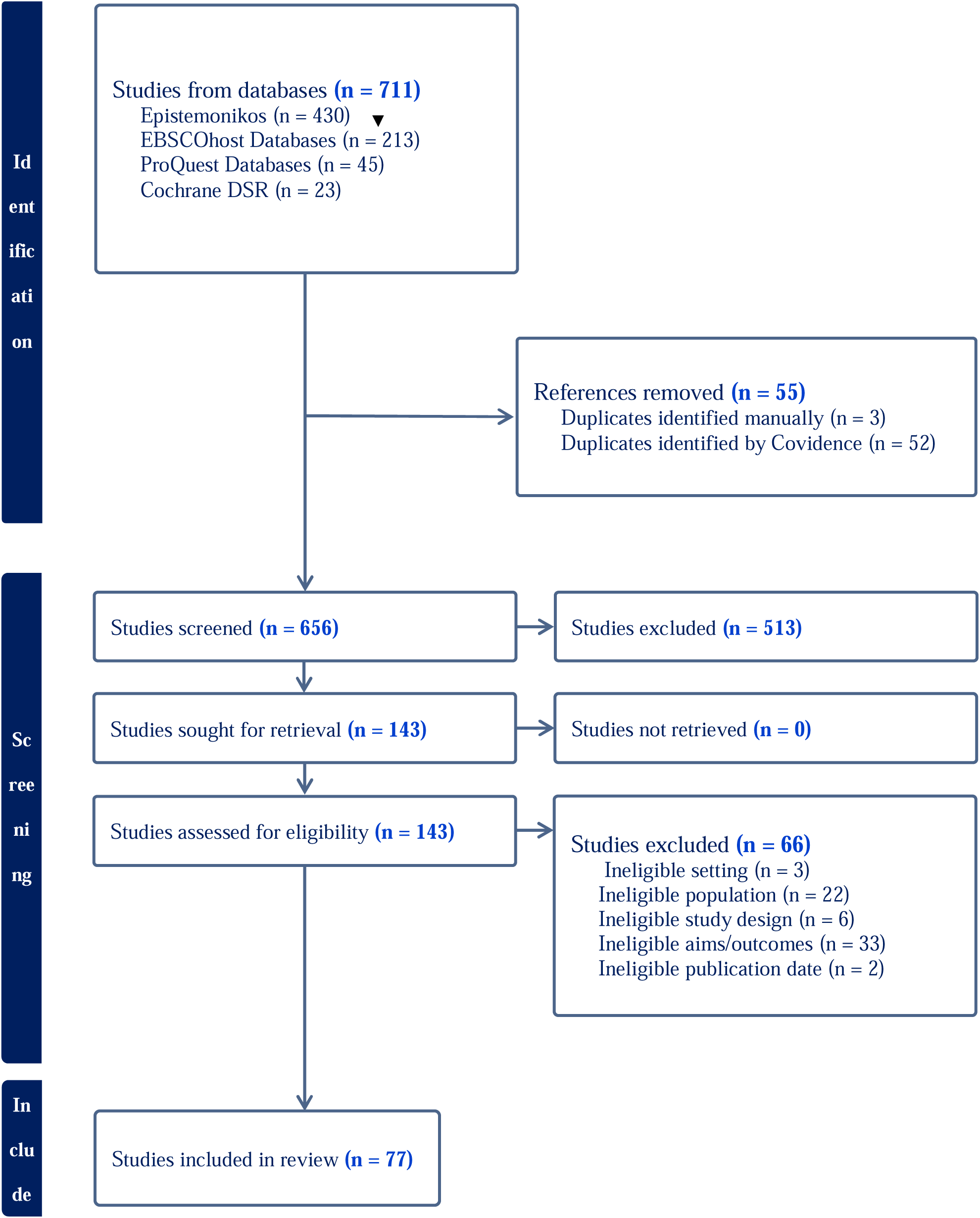
PRISMA flow diagram illustrating the study selection process

### Patterns in OHC outcomes reported in the included reviews

Publication activity on the outcomes of OHC increased steadily from 2013 (1 review) to a peak in 2018 and 2019 (11 reviews each). However, the trend declined in subsequent years, with only 5 reviews in 2023. The most frequently reported outcome domain was *Living Standards and Social Wellness* (LSSW), with 53 reviews reporting on such outcomes. The number of reviews on the *Health and Emotional Wellbeing* (HEW) domain remained relatively stable, with 4-6 reviews published annually since 2016. There was a noticeable rise in *Education and Learning* (EL) reviews around 2017, with modest levels afterward. *Identity and Civic Participation* (ICP) and *Physical and Legal Security* (PLS) domains were least reported. Full details of these trends are presented in Figure 2.

**Figure 2.**
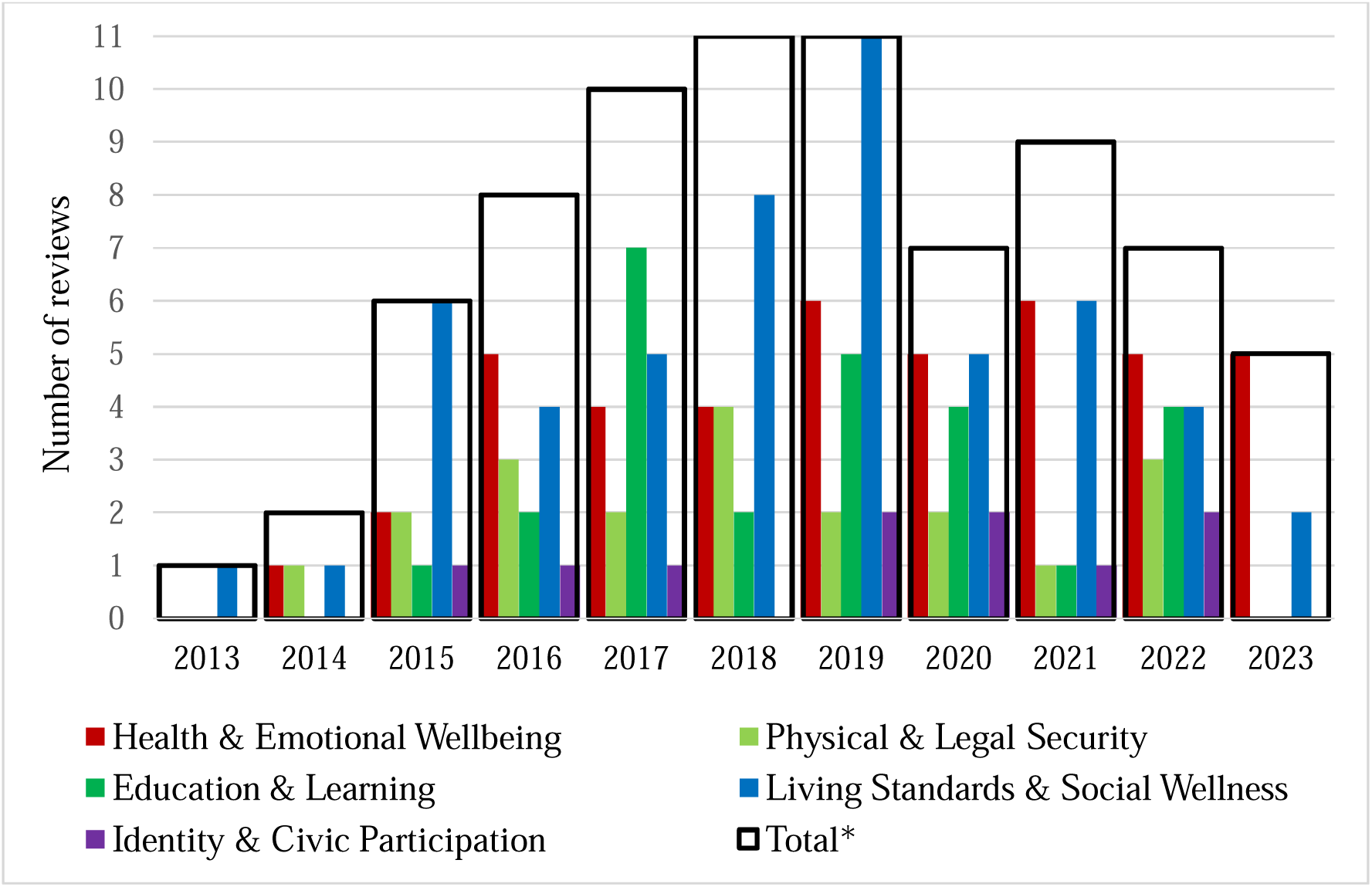
Trends in the number of review studies on outcomes for children in OHC (2013– 2023). *Might not equal the sum of the values for the within-year domains as reviews within the same year might cover more than one domain.

The reviews highlight patterns in research focus across outcome domains and indicators, with certain areas receiving more attention than others. Within *Health and Emotional Wellbeing* (HEW), *mental and emotional health* was the most explored indicator (28 reviews), while *all-cause mortality* was least reported (4 reviews). In the *Physical and Legal Security* (PLS) domain, *offending behaviours* was the most reported (11 reviews), whereas *victim of violent crime*, *general safety and risk* and *experienced bullying* were the least reported (2 reviews each). For *Education and Learning* (EL), the most reported was *educational attainment* (14 reviews), while *educational readiness and access* received minimal attention (3 reviews). In the *Living Standards and Social Wellness* (LSSW) domain, *attachment and behaviour functioning* was the most reported (30 reviews), with *housing and accommodation* as the least reported indicator in this domain (8 reviews). In the *Identity and Civic Participation* (ICP) domain, *identity and self-respect* was the more frequently reported indicator (9 reviews), compared to *participation and influence* (3 reviews). These findings likely show a concentration of research on specific areas such as mental health and attachment, while highlighting gaps in others including bullying, mortality, and educational readiness. Full details on these patterns are presented in Figure 3.

**Figure 3.**
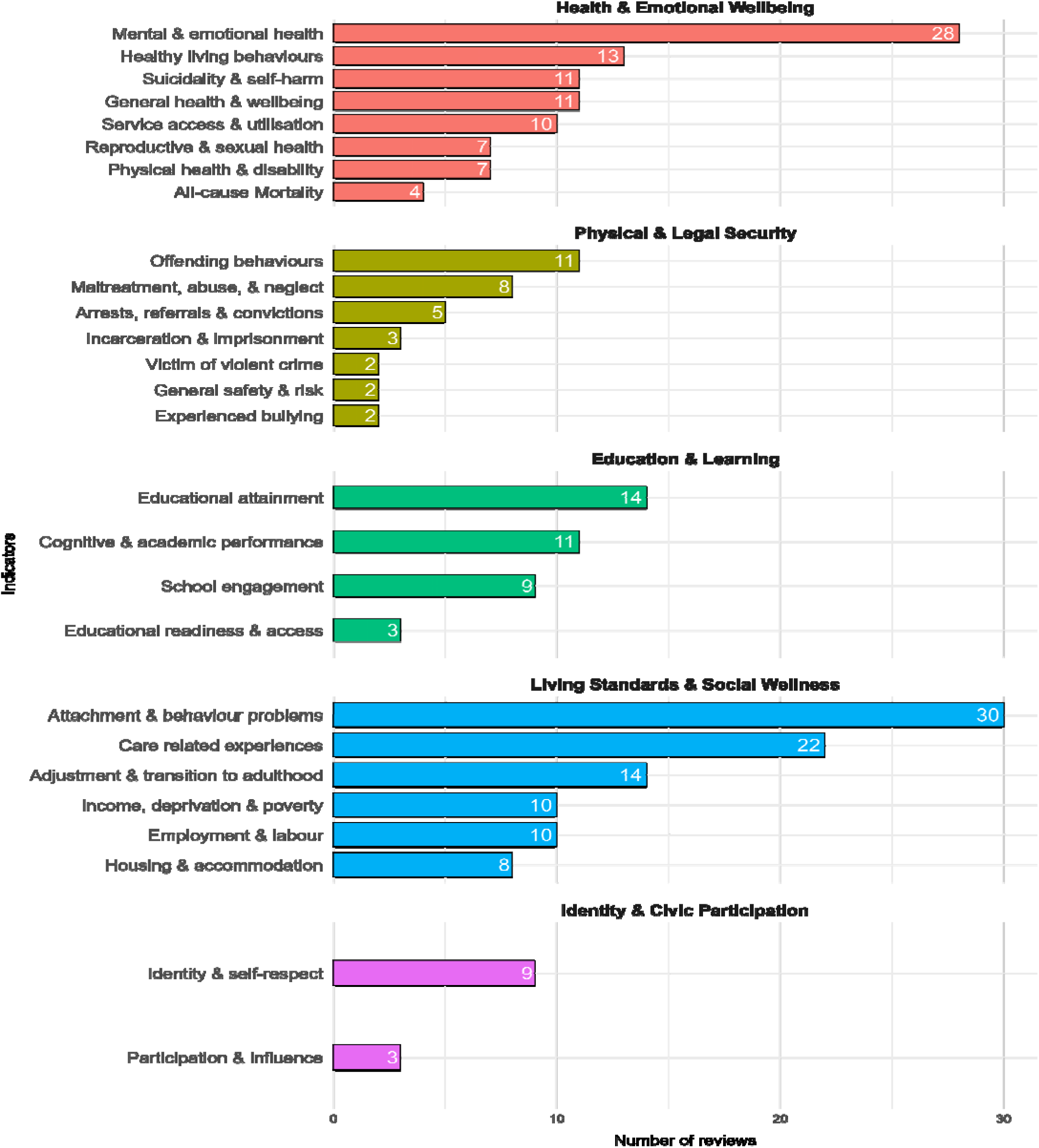
Number of review studies across outcome domains and indicators

### Factors associated with the outcomes of OHC

#### Factors reported across outcome domains

The most frequently reported factors related to HEW were at the system level (26 reviews), followed by the individual child level (21 reviews), and the socio-relational level (18 reviews). The most frequently reported factors related to PLS were at the socio-relational level (10 reviews), followed by the system level (9 reviews), and the individual child level (4 reviews). The most frequently reported factors related to EL were at the socio-relational level (15 reviews), followed by the system level (12 reviews), and individual child level (10 reviews). The most frequently reported factors related to LSSW were at the system level (30 reviews) and the socio-relational level (30 reviews), followed closely by the individual child level (25 reviews). The most frequently reported factors related to ICP were at the socio- relational level (4 reviews), and the other factors category (4 reviews), followed closely by the system level (3 reviews). Additional details on these patterns are shown in Figure 4.

**Figure 4.**
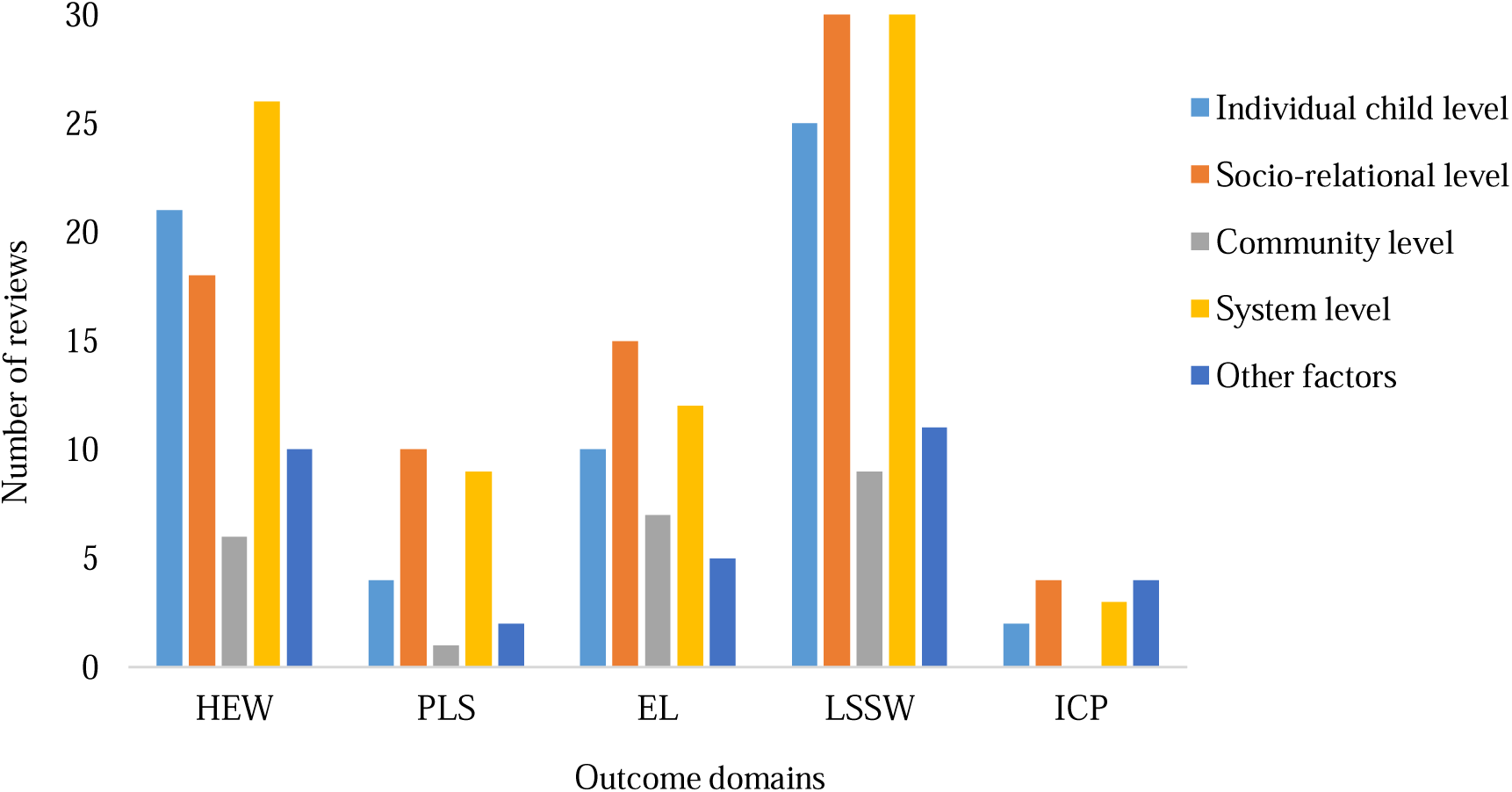
Frequency of factor reporting across outcome domains

### Health and Emotional Wellbeing

Within the HEW domain, varying number of reviews reported different factors in relation to the various outcome indicators. *Mental and emotional health* was most frequently reported in relation to system-level factors (16 reviews) and socio-relational factors (15 reviews), with community-level influences least explored (2 reviews). S*uicidality and self-harm* was commonly reported in relation to system-level factors (6 reviews), with no reviews reporting community-level influences. *All-cause mortality* was primarily reported in relation to individual child-level factors (3 reviews), while no review reported socio-relational or community-level influences. *Physical health and disability* was most reported in relation to system-level factors (3), whereas community-level factors were not reported. *Reproductive and sexual health* was often reported in relation to socio-relational factors (4), while community-level and other influences were least reported (1 review each). *Service access and utilisation* was also most frequently reported in relation to individual-level factors (5 reviews) and system-level factors (4 reviews), with community-level and other factors least reported (1 review each). For healthy living behaviours, individual child-level factors (6 reviews) and system-level factors (5 review) were the most frequently reported, whereas community-level factors were not reported. Finally, *general health and wellbeing* was commonly reported in relation to individual child-level factors (7 reviews), with community-level factors least reported (1 review). These patterns suggest that while system-level and individual-level factors dominate discussions on HEW indicators, community-level influences remain markedly underrepresented. Full details on these patterns are presented in Figure 5.

**Figure 5.**
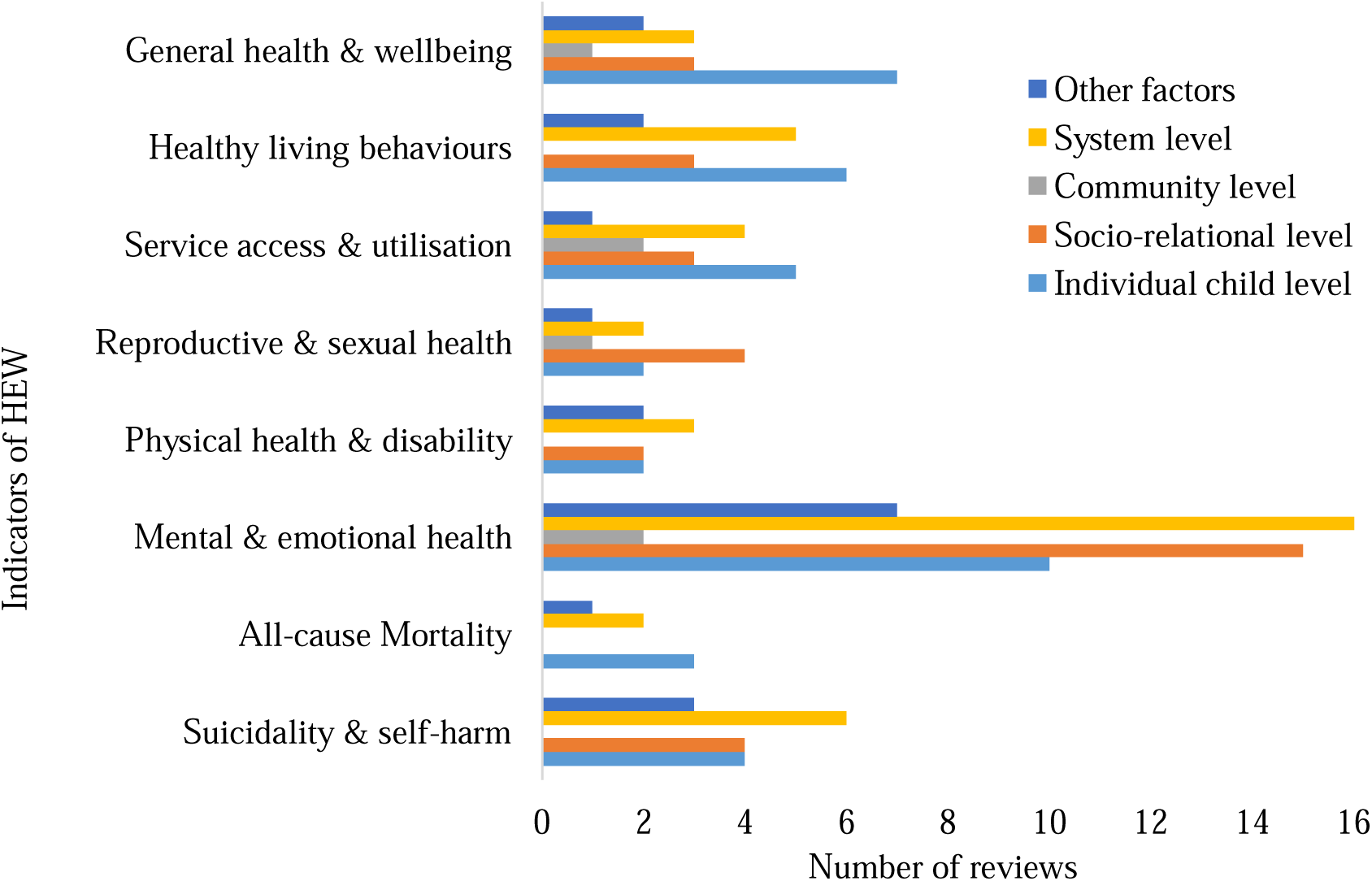
Distribution of HEW indicators across factor level

### Physical and legal security

In the PLS domain, at least one factor level was absent from the reporting of every outcome indicator. Notably, no outcome indicator was reported in relation to community-level factors, with the sole exception of *arrests, referrals and convictions*. Becoming a *v*ictim *of violent crime* was most frequently reported in relation to individual child-level (2 reviews). *Maltreatment, abuse, and neglect* was often reported in relation to socio-relational factors (3), followed by individual-level and system level factors (2 reviews each). *General safety and risk* was reported in relation to only system-level factors (3 reviews) and socio-relational factors (1 review). Two reviews reported *experienced bullying* in relation to system-level factors. *Arrests, referrals and convictions* was most reported in relation to system-level and socio-relational factors (2 reviews each). *Incarceration and imprisonment* was reported in relation to only system-level factors (2 reviews) and socio-relational factors (1 review). Offending behaviours was most frequently reported in relation to system-level factors (6 reviews), followed by socio-relational factors (5 reviews). Full details on these patterns are presented in Figure 6.

**Figure 6.**
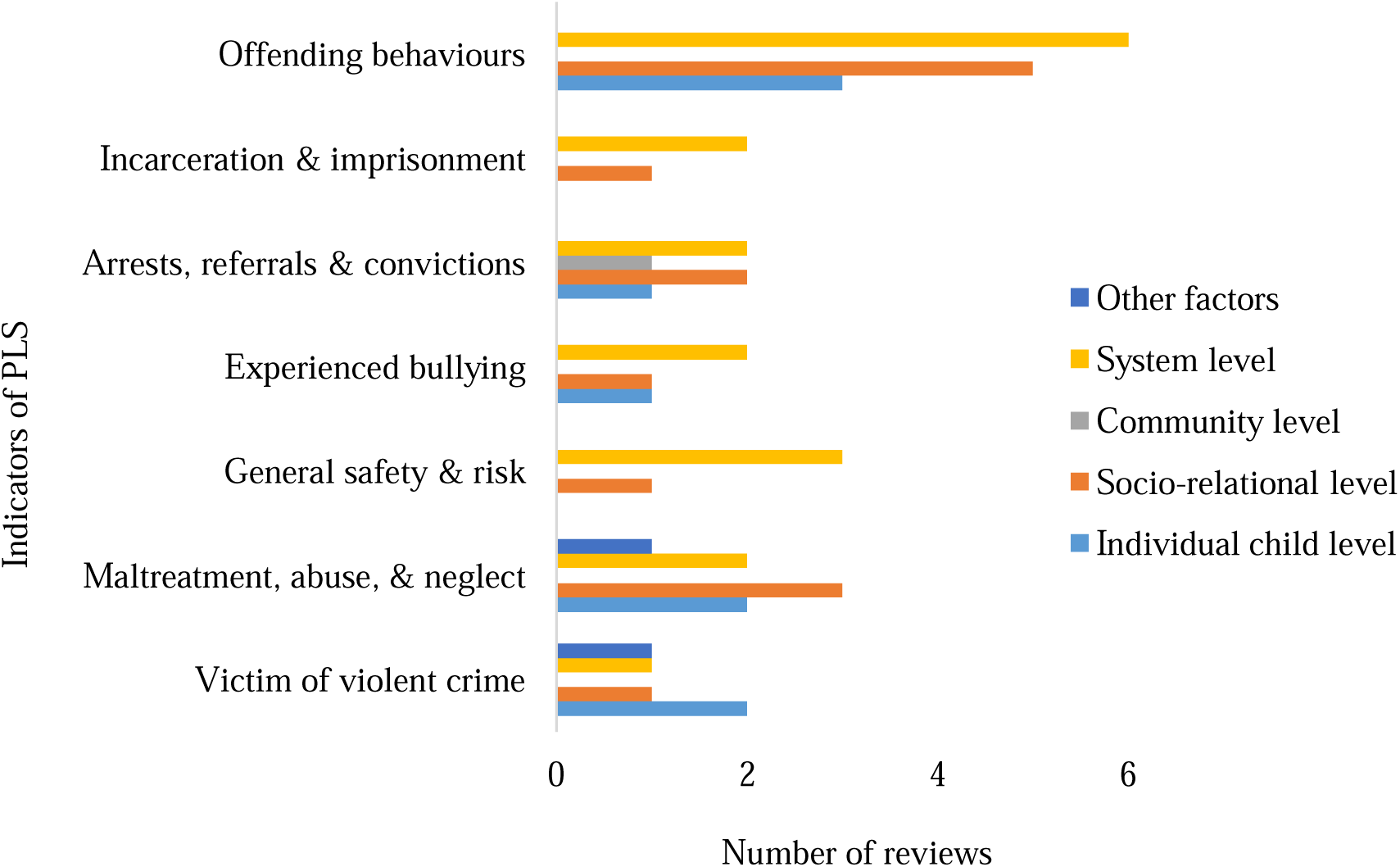
Distribution of PLS indicators across factor levels

### Education and learning

Within the EL domain, all outcome indicators were reported across all factor levels, with the exception of *educational readiness and access* which was not reported in relation to other factors. *Educational attainment* was most frequently reported in relation to individual child-level factors (8 reviews), followed by socio-relational factors (6 reviews), and with other factors less commonly reported (3). *Cognitive and academic performance* was most frequently reported in relation to individual child-level and system-level factors (5 reviews each), while community-level influences were least reported (1 review). *School engagement* was most often reported in relation to system-level factors (6 reviews), followed by socio- relational factors (5 reviews), and other factors were least reported (1 review). *Educational readiness and access* was evenly reported in relation to system, socio-relational, and individual child-level factors (2 reviews each). Full details on these patterns are presented in Figure 7.

**Figure 7.**
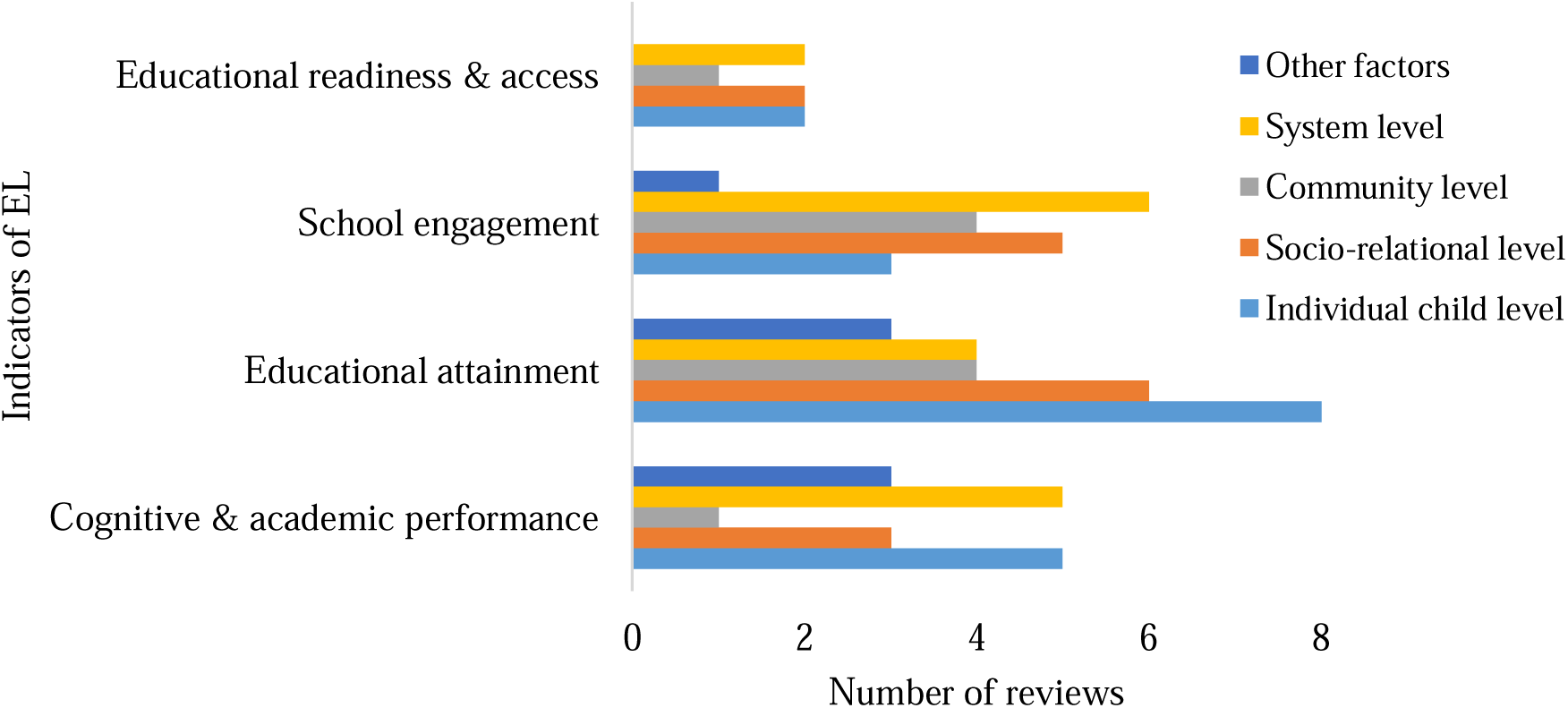
Distribution of EL indicators across factor levels

### Living standards and social wellness

Within the LSSW domain, all outcome indicators were reported across all factor levels, except *income, deprivation and poverty* which was not reported in relation to community-level and other factors. *Attachment and behaviour functioning* was most frequently reported in relation to system-level (18 reviews) and socio-relational factors (16 reviews), while community-level influences were less commonly reported (4 review). *Income, deprivation and poverty* was most frequently reported in relation to system-level factors (6 reviews) and individual child-level factors (4 reviews). *Care related experiences* was most often reported in relation to socio-relational factors (14 reviews), with community-level and other factors least reported (2 reviews each). *Employment and labour* was most commonly reported in relation to individual child-level factors (5 reviews) and system-level factors (4 reviews), with community-level factors least reported (1 review). *Adjustment and out of care issues* was equally reported in relation to system-level and socio-relational factors (6 reviews each), with less reporting of community-level and other factors (2 reviews each). *Housing and accommodation* was most frequently reported in relation to system-level factors (4 reviews), with community-level and other factors least reported (1 review). Full details on these patterns are presented in Figure 8.

**Figure 8.**
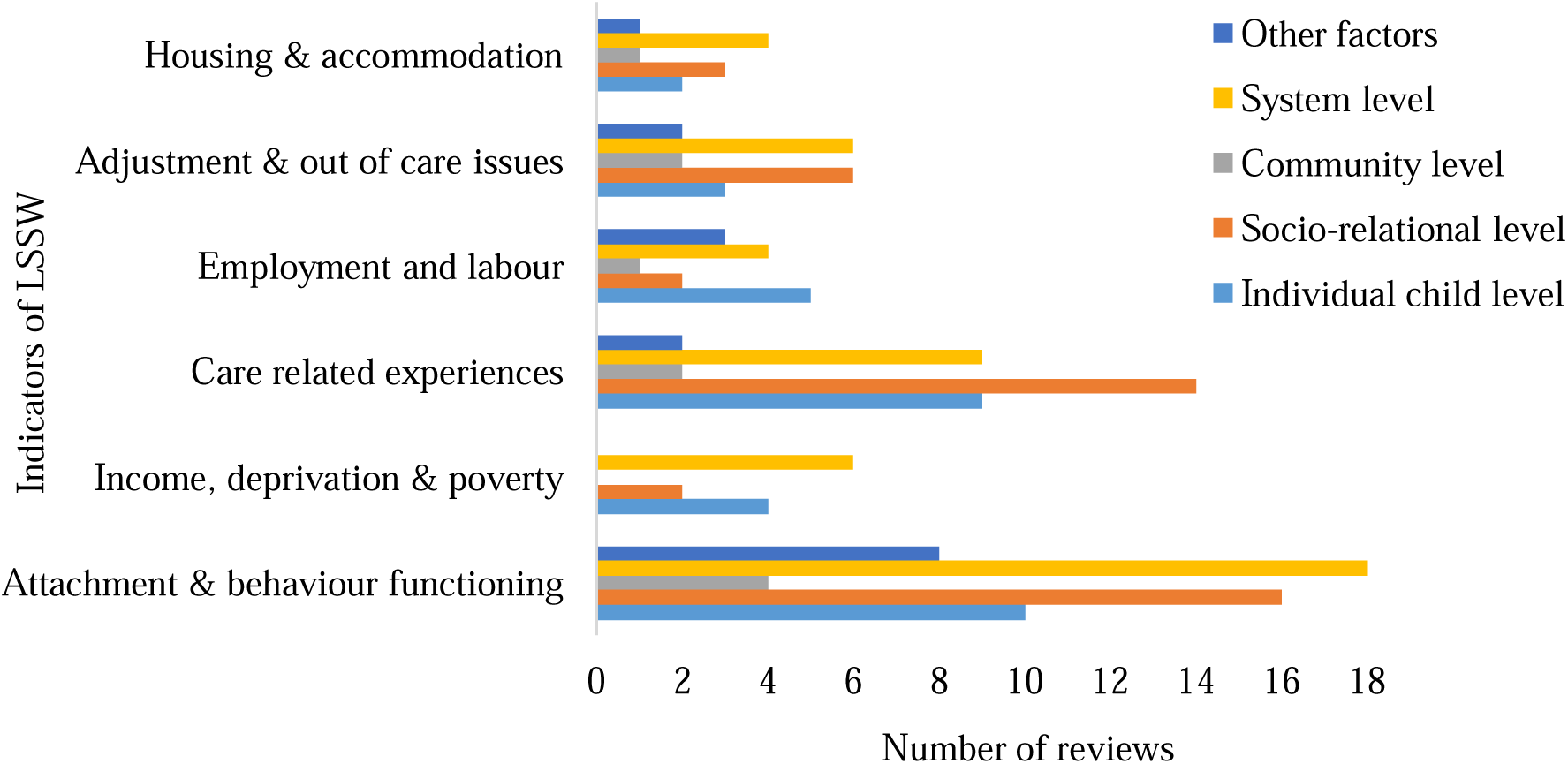
Distribution of LSSW indicators across factor levels

### Identity and Civic Participation

Community-level factors were not reported in the reviews covering ICP outcome indicators. *Identity and self-respect* was most frequently reported in relation to socio-relational and other factors (4 reviews each), while system-level and individual child-level factors were less commonly reported (2 reviews each). *Participation and influence* was only reported in relation to other factors (2 reviews) and system-level factors (1 review). Full details on these patterns are presented in Figure 9.

**Figure 9.**
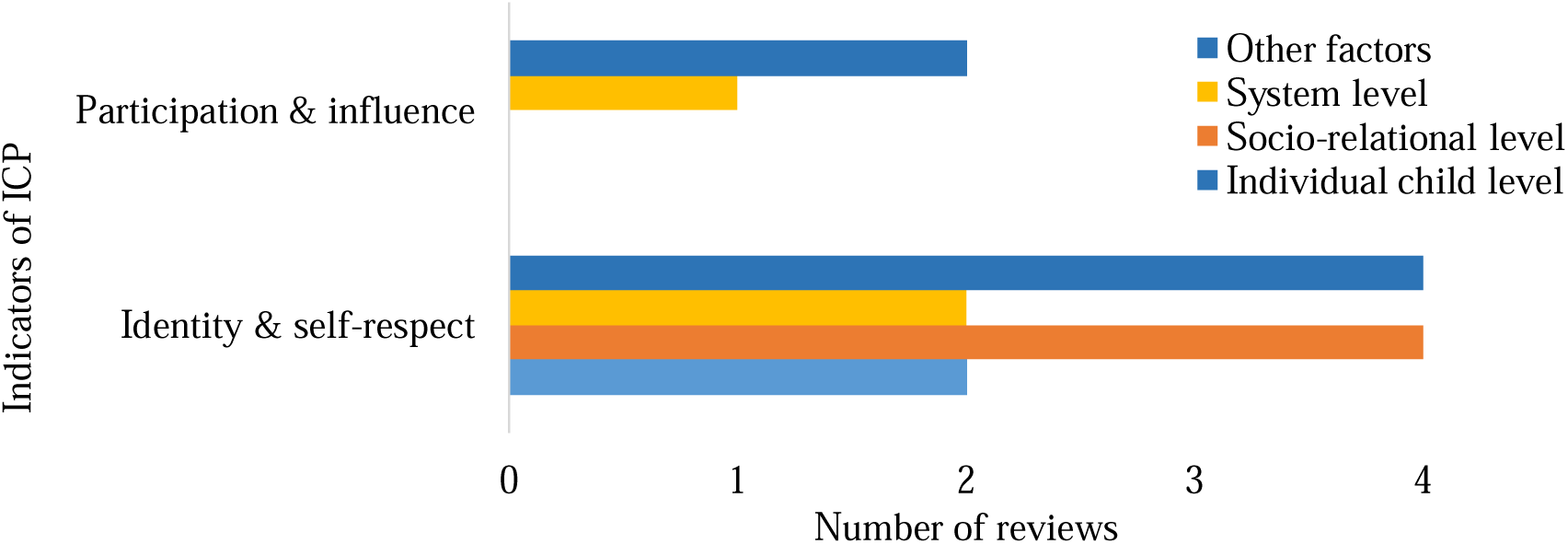
Distribution of identity and civic participation indicators across factor levels

## Discussion

This scoping review of reviews included 77 studies to systematically map existing reviews on the outcomes of OHC and the factors examined in relation to these outcomes. The results underscore critical patterns across domains of HEW, PLS, EL, LSSW, and ICP, offering valuable insights into the factors reported in relation to these outcomes with the potential to inform future efforts.

### Patterns and gaps in research focus

This evidence map demonstrates a concentration of research efforts on specific of care-experienced individuals, particularly *Living Standards and Social Wellness* (LSSW) and *Health and Emotional Wellbeing* (HEW), both of which are closely tied to children’s immediate and long-term quality of life [13,34]. The consistent volume of research in these areas may reflect their prioritisation in policy and practice. However, *Identity and Civic Participation* (ICP) emerged as the least reported domain, with far fewer reviews addressing outcomes about identity development, self-respect, influence and community participation. This disparity might suggest a critical gap in primary research, particularly given the importance of identity development and civic participation in fostering a sense of belonging and resilience among care-experienced children and young people [15]. Research on indicators often reflected uneven attention across domains. For example, within *Health and Emotional Wellbeing* (HEW), substantial emphasis was found on mental and emotional health, while indicators such as all-cause mortality had received little attention. A similar pattern was observed in *Physical and Legal Security* (PLS), where offending behaviours were well documented, whereas indicators such as bullying and being a victim of violent crime were relatively underrepresented. These patterns suggest a need for more balanced research efforts to ensure that less reported but equally critical aspects of children’s experiences are adequately addressed.

The review highlighted the prominence of system level factors (e.g. placement types and care quality) across all outcome domains. These factors were most frequently reported in relation to indicators such as attachment and behaviour functioning, as well as mental and emotional health. This finding aligns with a previous correlates review [96], highlighting the significant role of system and carer-related factors, particularly their impact on placement stability and behaviour of children in out-of-home care (OHC). However, while system-level factors were well-represented, community factors were minimally reported across all outcome domains. This gap may point to an unmet need for research exploring the influence of community-level factors such as neighbourhood characteristics in influencing outcomes of children in OHC.

The findings of this review of reviews also highlight the consistent reporting of socio- relational factors across several outcome indicators. Social relationships—such as those with family and peers—often serve as protective factors, highlighting the importance of maintaining familial connections where possible and fostering meaningful relationships for care-experienced children [41,53,58,94]. Nonetheless, the underrepresentation of socio- relational factors in domains such as *Identity and Civic Participation* (ICP) and *Legal and Physical Security* (PLS) points to potential areas for further exploration.

### Strengths and limitations

This review represents a comprehensive analysis of review-level evidence, providing an overview of outcomes and associated factors for children in OHC. By extracting data from full texts rather than just abstracts, this study minimised the risk of missing relevant outcomes and factors. The involvement of care-experienced people in the study strengthened the review by incorporating diverse perspectives in refining the search strategy and contextualising findings [19]. However, limitations include the variability in methodological approaches and scope across the included reviews, which may affect the generalisability of some findings. The reliance on descriptive analysis, while useful for mapping the evidence base, limits the depth of the review’s conclusions. Nonetheless, the data and specific factor-outcome relationships are provided as a resource for stakeholders seeking evidence on factors influencing outcomes for children in OHC (see S4 File). It is worth noting that some included reviews [42,64,69,73,76] did not report moderating variables influencing outcomes but focused solely on outcomes for children following placement in OHC. These reviews typically compared children in OHC with those in the general population, offering limited insight into factors shaping various outcomes within the care-experienced group.

### Future directions

Future research should aim to address the identified gaps by focusing on underexplored domains, such as *Identity and Civic Participation* (ICP) and indicators including bullying, mortality, and educational readiness. Greater attention should be given to community level factors, as these are often underexplored despite their vital role in supporting children’s transitions to independence and social integration. Methodologies such as participatory research, involving care-experienced children and young people, can provide deeper insights into their lived experiences and priorities. Policymakers, practitioners, and researchers must collaborate to ensure that the evidence generated is translated into actionable policies and practices that improve outcomes for all children in OHC.

## Conclusion

This review of reviews highlights significant patterns and gaps in the research on outcomes for children in OHC. *Living Standards and Social Wellness* (LSSW), along with *Health and Emotional Wellbeing* (HEW), have been most reported in the literature, perhaps reflecting their relative importance. In contrast, domains such as *Physical and Legal Security* (PLS) and *Identity and Civic Participation* (ICP) remain underexplored, underscoring the potential need for greater research in these areas. System-level factors dominate the discourse, particularly in shaping behavioural functioning, health and educational outcomes, while community-level influences remain underrepresented.

## Data Availability

All relevant data are within the manuscript and its Supporting Information files.

## Acknowledgements

We would like to express our gratitude to the Welsh National Centre for Population Health and Wellbeing Research PPI group for their invaluable contributions during the grant development and submission process, which informed the PPI strategy for the project. We also extend our heartfelt thanks to CASCADE Voices, a partnership of the Children’s Social Care Research and Development Centre (CASCADE) and Voices from Care Cymru, for facilitating access to care-experienced children, young people, and parents involved with child welfare during the planning and execution of this review of reviews. CASCADE receives infrastructure funding from Health and Care Research Wales.

## Author contributions

**Richmond Opoku:** Conceptualization, Methodology, Data curation, Formal analysis, Writing – original draft, Writing – review & editing; **Natasha Judd:** Conceptualization, Methodology, Data curation, Formal analysis, Validation, Writing – review & editing; **Katie Cresswell:** Conceptualization, Methodology, Data curation, Formal analysis, Writing – review & editing; **Michael Parker:** Conceptualization, Methodology, Data curation, Formal analysis, Writing – review & editing; **Michaela James:** Conceptualization, Methodology, Data curation, Validation, Writing – review & editing, Supervision; **Jonathan Scourfield:** Conceptualization, Methodology, Writing – review & editing, Supervision; **Karen Hughes:** Conceptualization, Methodology, Formal analysis, Validation, Writing – review & editing, Supervision; **Jane Noyes:** Conceptualization, Methodology, Writing – review & editing, Supervision; **Dan Bristow:** Conceptualization, Methodology, Writing – review & editing, Supervision; **Evangelos Kontopantelis:** Conceptualization, Methodology, Data Curation, Writing – review & editing, Supervision. **Sinead Brophy:** Conceptualization, Methodology, Data curation, Validation, Writing – review & editing, Supervision; **Natasha Kennedy:** Conceptualization, Methodology, Data curation, Formal analysis, Writing – review & editing, Supervision.

