## Supplementary material for "Mapping the Evidence on Outcomes of Childhood Out-of-Home Care: A Scoping Review of Reviews": S2 Table

**Article Title: Mapping the Evidence on Outcomes for Children in Out-of-Home Care: A Scoping Review of Reviews**

**S1 Table. Development of outcome domains and indicators**

| **Domain/Indicator** | **Code in Data** | **Source** | **EMF Domain(s)** | **EMF Indicator(s)** | **Remarks** |
| --- | --- | --- | --- | --- | --- |
| **Health & emotional wellbeing** | 1 | EMF | A and B | N/A | Combined the two related EMF domains to create a single domain |
| Suicidality & self-harm | a3 | Both | A | 3 | Indicator broadened to include suicidal ideation, plans, attempts, and self-harm due to sparse reporting, making separate indicators impractical. |
| All-cause Mortality | a5 | Data | A | 5 | Not immediately available in EMF. Association to EMF indicator is mainly for easy categorisation and analysis. |
| Mental & emotional health | b1m | EMF | B | 1 | Considered together with physical health and disability in EMF |
| Physical health & disability | b1p | EMF | B | 1 | Considered together with mental and emotional in EMF |
| Reproductive & sexual health | b2 | EMF | B | 2 |  |
| Service access & utilisation | b3 | EMF | B | 3 |  |
| Healthy living behaviours | b4 | EMF | B | 4 |  |
| General health & wellbeing | b5 | Both | B | 5 | Indicator broadened to include generally defined health and wellbeing indicators in the data (e.g. health status, wellbeing, quality of life etc) |
| **Physical & legal security** | 2 | EMF | C and D | N/A | Combined the two related EMF domains to create a single domain |
| Victim of violent crime | c1 | EMF | C | 1 |  |
| Maltreatment, abuse, & neglect | c2 | EMF | C | 2 |  |
| General safety & risk | c5 | Both | C | 5 | Indicator name modified to reflect the data |
| Experiences of bullying | c6 | EMF | C | 6 |  |
| Arrests, referrals & convictions | d1 | Both | D | 1 | Indicator broadened to include arrests, referrals and convictions. |
| Incarceration & imprisonment | d2 | Both | D | 2 | Indicator broadened to include incarceration and imprisonment. |
| Offending behaviours | d5 | Data | D | 5 | Indicator broadened to include all offending behaviours pepetrated by children in care as EMF mainly focuses on children as victims. Association to EMF indicator is mainly for easy categorisation and analysis. |
| **Education & learning** | 3 | EMF | E | N/A |  |
| Academic performance | e1 | EMF | E | 1 | Indicator name modified to reflect the data |
| Educational attainment | e2 | Data | E | N/A | No practical associations with EMF indicators |
| School engagement | e3 | Data | E | N/A | No practical associations with EMF indicators |
| Educational readiness & access | e4 | Data | E | N/A | No practical associations with EMF indicators |
| Living standards & social wellness | 4 | EMF | F,G and H | N/A | Combined the three related EMF domains to create a single domain |
| Housing & accommodation | f1 | EMF | F | 1 |  |
| Income, deprivation & poverty | f2 | EMF | F | 2 |  |
| Care related experiences | f3 | Both | F | 5 | Indicator broadened to include care related outcomes (e.g., placement instability, length of placement etc.) |
| Employment and labour | g3 | EMF | G | 3 and 4 | Combined the two related EMF indicators to create a single indicator |
| Adjustment & out of care issues | h0 | Data | H | N/A | No practical associations with EMF indicators |
| Attachment & behaviour problems | h3 | Both | H | 3 | Indicator broadened to include other behaviour problems (e.g., externalising and internalising behaviours etc.) |
| Identity & civic participation | 5 |  | I and J | N/A | Combined the two related EMF domains to create a single domain |
| Identity & self-respect | i | EMF | I | 1-5 | All EMF indicators were consolidated into one indicator due limited data availability in this domain |
| Participation & influence | j | EMF | J | 1-5 | All EMF indicators were consolidated into one indicator due limited data availability in this domain |

EMF = Equality Measurement Framework; A = Life; B = Health; C = Physical Security; D = Legal Security; E = Education and Learning; F = Standard of Living; G = Productive and Valued Activities; H = Individual, Family and Social Life; I = Identity, Expression and Self-respect; J = Participation, Influence and Voice
