## Supplementary material for "Mapping the Evidence on Outcomes of Childhood Out-of-Home Care: A Scoping Review of Reviews": S3 Table

### Article Title: Mapping the Evidence on Outcomes for Children in Out-of-Home Care: A Scoping Review of Reviews

### S2 Table. Study characteristics

| **Author, year** | **Type of review** | **Number of studies included (*)** | **Type of included studies** | **Location of included studies** | **Review time frame** | **Population included** |
| --- | --- | --- | --- | --- | --- | --- |
| Welch, 2015 | Selective Review | 90 (90) | Qa, Q1, Md | U.S.A = 54, U.K. = 20, Canada = 10, Australia = 4, China =1 and The Netherlands =1 | 1998 – 2013 | Children and young people in foster care and adoption |
| Geiger, 2017 | Scoping Review | 51(51) | Q1, Qa & Md | US = 50, Europe = 1 | 1997-2015 | Foster care alumni in post-secondary education |
| Goding, 2022 | Systematic Review | 11(11) | Qa & Md | UK | 2010-2019 | Children and young people in care |
| Batty, 2022 | Systematic Review & Meta-analysis | 14 (14) | Q1 | Finland = 4, Sweden = 4, UK = 3, Canada = 1, USA = 1, and Australia = 1 | 1995-2022 | Children (up to 18 years) exposed to temporary out-of-home care |
| Phillips, 2023 | Scoping Review | 23(23) | Q1, Qa | USA = 11, UK = 3, Australia = 2, Canada = 2, Israel = 2, Spain = 1, and Sweden = 1 | 2006 - 2021 | Care-leavers, defined as any person who was under state care when they crossed the age of eligibility for being under state care. |
| Zabern, 2018 | Literature Review | 17 (17) | Q1, Qa, Md | n.r | 2003 - 2016 | Children in out-of-home care. |
| Washington, 2018 | Systematic Review | 40 (40) | Q1 | USA = 40 | 2010 - 2016 | Children residing in foster or kinship care. |
| Yoon, 2018 | Systematic Review | 15 (15) | Q1 | n.r | 1985 - 2017 | Current or former youth in out-of-home care. |
| Tang, 2023 | Systematic Review | 13 (6) | Q1 | USA = 4, Finland = 4, UK = 3, Australia = 1, and Luxembourg = 1. | 2009 - 2022 | Children <18 years of age exposed to early institutionalization, placement into foster care, parental incarceration, separation when parents migrate for economic reasons, separation due to asylum and war. |
| Kääriälä, 2017 | Systematic Review | 20 (20) | Q1 | Sweden = 12, Finland = 5, Norway = 2, and Denmark = 1 | 2001 - 2016 | Populations born in 1965 or later in Denmark, Finland, Norway, or Sweden; reported any outcome of OoHC beginning at age 18 or later; compared a population that was placed in OoHC with a population that was never in OoHC during their childhood or adolescence. |
| Day, 2018 | Systematic Review | 48 (48) | Qa | n.r | 1987-2016 | Resource parents who care for older youth |
| Häggman-Laitila, 2020 | Integrative review | 16 (16) | Q1, Qa | USA = 11, Australia = 2, Finland = 1, Israel = 1, UK = 1 | 2011 - 2018 | Care leavers who have left the foster care |
| O'Higgins, 2017 | Systematic Review | 39(39) | Q1 | USA = 24, Canada = 5, UK = 5, Australia = 4 and Sweden = 1 | 1992 - 2014 | School-age children in foster or kinship care. |
| Cassarino-Perez, 2018 | Meta-analysis | 24 (24) | Q1 | USA = 14, UK = 1, Spain = 1, Sweden = 1 | 2006 - 2015 | Young people aging out of care (between 16 and 30 years old) |
| Quarmby, 2016 | Scoping Review | 7(7) | Q1, Qa | USA =3 , UK = 2 and Norway 2 | 1998 - 2013 | Children and young people living in or leaving care |
| Häggman-Laitila, 2018 | Systematic Review | 21 (21) | Qa | UK = 5, USA = 4, Canada = 2, Ghana = 2, India = 1, Ireland = 1, Norway = 1, Republic of Korea = 1, Romania = 1, South Africa = 1, Sweden = 1, Zimbabwe = 1 | 2010 - 2017 | Young people leaving foster care |
| Doab, 2015 | Systematic Review | 11(11) | Q1, Qa | USA | 2007-2012 | Mothers with Substance Use Disorders (SUD) |
| Strijbosch, 2015 | Meta-analysis | 19 (19) | Q1 | North America = 15, Western Europe = 4 | 1987 - 2012 | Children and youth between 4 and 17 years old who are in care |
| Hurry, 2023 | Scoping Review | 28(28) | Q1 | UK = 28 | 2001-2021 | Looked after children up to the age of 18 (extended to 22 years if the LAC was still in full time education) |
| Loomis, 2020 | Systematic Review | 18(12) | Q1 | USA = 18 | 1999-2017 | Older youth currently or formerly in foster care (13 years+) |
| Townsend, 2020 | Systematic Review | 11 (11) | Qa, Md | USA = 7, UK = 2, Canada = 1, Germany = 1 | 2003 - 2018 | Children currently in care and youth/adults who had previously been in care. |
| Winokur, 2018 | Systematic Review | 102(102) | Q1, Qa | USA = 89 | 1991 - 2011. | Children and youth less than 18 years who were removed from the home for abuse, neglect, or other maltreatment and subsequently placed in kinship care. |
| Johnson, 2019 | Systematic Review | 46(46) | Qa, Q1, Md | USA = 46 | 1990 - 2018 | Undergraduate Youth formerly in foster care (YFFC) |
| Bronsard, 2016 | Systematic Review & Meta-analysis | 8 (8) | Q1 | UK = 3, USA = 2, France = 1, Germany = 1, Norway = 1 | 1996 - 2013 | Children and adolescents involved in the child welfare system |
| Goemans, 2015 | Meta-analysis | 29 (29) | Q1 | USA = 15, The Netherlands = 4, Australia = 2, Croatia = 1, Iraq = 1, Norway = 1, Northern Ireland = 1, Scotland = 1, Canada = 1, UK = 1, Belgium = 1 | 1978 - 2013 | Children in regular foster care aged 0-18 years |
| Randolph, 2017 | Systematic Review | 7 (4) | Q1, Qa, Md | USA = 7 | 2008 - 2015 | Care experienced youth |
| Fry, 2017 | Systematic Review | 31 (3) | Q1 | USA = 18, South America = 4, Canada = 2, Sweden = 2, Israel = 2, UK = 1, Caribbean = 1, Seychelles = 1 | 1995 - 2014 | Young people who have experienced foster care |
| Costa, 2019 | Systematic Review | 25 (25) | Q1 | Spain = 4, Portugal = 7, Israel = 3, Brazil = 2, USA = 1, France = 1, UK = 1, Japan = 1, Iran = 1, Estonia = 1, Korea = 1, Croatia = 1, Romania = 1, | 2000 - 2017 | Adolescents (aged 11-18 years) in residential or institutional care |
| Carr, 2020 | Systematic Review | 49(49) | Md | Austria =12, Switzerland =9, Canada =6, Ireland =5, USA =3, Australia =2, The Netherlands =1, Germany =1 | 1996-2018 | Children in long-term care |
| Engler, 2020 | Systematic Review | 25(25) | Q1 | USA =18, Norway =3, UK =1, Germany =1, Canada =1, New Zealand =1 | 2001-2018 | Children who have been in foster care |
| Lou, 2018 | Systematic Review | 15(15) | Q1, Qa, Md | USA =2, Singapore =3, South Africa =2, Canada =1, Israel =1, Croatia =1, Portugal =1, Iran =1, Czech Republic =1, Greece =1 | 2002-2017 | Youth (<19 years) in residential care settings |
| DiGiovanni, 2021 | Literature Review | 43 (43) | Q1, Qa, other - reviews | Reported for some studies (includes USA, Australia, North Pacific region) | 1960 - 2019 | Children in foster care |
| DeLuca Bishop, 2019 | Meta-analysis | 37(7) | Q1 | USA =17, Spain =2, France =1, Israel =1 | 1983-2015 | Those with foster care experience |
| Collins, 2016 | Integrative review | 18 (18) | Q1, Qa | n.r | 2006 - 2015 | Youth (12 to 30 years of age) in foster care or transitioning out of foster care |
| Wilson, 2020 | Systematic Review | 12 (12) | Q1 | USA = 3, India = 2, Australia = 1, Haiti = 1, Israel = 1, Portugal = 1, Spain = 1, Turkey = 1, Russia = 1 | 1989 - 2019 | Children or adolescents (under the age of 20) and drawn from a full-time care setting, with admission due to abuse or neglect at home, parental incapacity, death or inability to provide for the child's needs. |
| Zhang, 2021 | Meta-analysis | 15 (15) | Q1 | USA = 13, Ireland = 1 and Switzerland = 1. | 2012 - 2020 | Children involved with the child welfare system (including children adopted from the child welfare system). |
| Xu, 2018 | Systematic Review | 8 (8) | Q1 | USA = 6, Belgium = 1, Norway = 1. | 2012 - 2017 | Children or adolescents in kinship or non-kinship foster care |
| Evans, 2017 | Systematic Review & Meta-analysis | 5 (5) | Q1 | Canada = 2, USA = 1, England = 1, Australia = 1 | 2001 - 2011 | Children and young people that have been placed in care |
| Cameron-Mathiassen, 2022 | Systematic Review | 12(12) | Qa | UK =6, Sweden =2, Norway =1, USA =1, South Africa =1, Australia =1 | 2002-2019 | Children, youths and young adults within the age range 12 to 25 years, living in residential institutional care of a continuous or ongoing nature. Young people whose residency was not related to physical or learning disabilities |
| Lee, 2023 | Systematic Review | 24(24) | Q1 | USA = 24 | 1994-2020 | Children and youth in the USA foster system |
| Thompson, 2016 | Systematic Review | 38 (23) | Q1, Qa, Md | USA = 23, n.r | 2006 - 2015 | Older youth in foster care. |
| Hayes, 2023 | Systematic Review | 20 (20) | Qa | UK = 9, USA = 3, The Netherlands = 2, Canada = 2, South Africa = 1, Denmark = 1, Ireland = 1 and Sweden = 1 | 2003 - 2021 | Children and young people currently living in foster care |
| Kang-Yi, 2017 | Systematic Review | 28 (28) | Q1, Qa, Md | n.r | 1991 - 2013 | Youth with behavioral health disorders aging out of foster care |
| Konijn, 2019 | Meta-analysis | 42(42) | Q1 | USA/Canada =24, Europe =16, Australia =2 | 1990-2015 | Long-term foster care in Western countries |
| Dubois-Comtois, 2021 | Meta-analysis | 41 (41) | Q1 | USA = 26, England = 3, Belgium = 2, Australia = 1, Canada = 1, Chile = 1, Croatia = 1, Ireland = 1, The Netherlands = 1, Norway = 1, Serbia = 1, Spain = 1, Turkey = 1 | 1988 - 2017 | Children living in foster care at the time of intake, with more than 75% of the children under 21 years of age. |
| Geiger, 2017 | Literature Review | 22 (22) | Q1, Qa, Md | n.r | 2005 - 2015 | Foster care alumni in higher education |
| Kekoni, 2019 | Integrative literature | 41(41) | Q1, Qa | n.r | 1993-2014 | n.r |
| Li, 2017 | Meta-analysis | 23 (23) | Q1 | USA = 10, UK = 2, Serbia = 2, Iraqi Kurdistan = 1, Croatia = 1, Canada = 1, The Netherlands = 1, Australia = 1, Romania = 1, South Korea = 1, Germany = 1, Singapore = 1 | 1999 - 2016 | Children in residential care compared to children in family foster care |
| BergstroÂ¨m, 2019 | Systematic Review & Meta-analysis | 28 (28) | Q1 | USA = 20, The Netherlands = 2, UK = 1 | 1994 - 2017 | Children up to the age of 17 who are placed in foster family care |
| Biehal, 2014 | Literature Review | 19 (18) | Q1 | USA = 11, UK = 7, Australia = 1 | 1981 - 2008 | Children in foster care |
| Mazzone, 2018 | Literature Review | 31(31) | Q1, Qa | Israel =7, England =5, Spain =2, Croatia =7, Japan =1, The Netherlands =1, Korea =1, Australia =1, n.r =6 | 1997-2017 | Children living in institutional care settings |
| Miranda, 2019 | Literature Review | n.r | n.r | n.r | n.r | Children in foster care and foster care alumni |
| Lund, 2020 | Scoping Review | 25(25) | Q1, Qa, Md | Australia =16, UK =3, International =3, New Zealand =2, Europe =1 | 2010-2020 | Children and young people in out-of-home care in Australia |
| Ãlvarez, 2022 | Scoping review | 14 (14) | Q1, Qa, Md | USA = 14 | 2002-2019 | LGBTQIA+ children and youth in out-of-home care |
| Quiroga, 2016 | Systematic Review | 18 (18) | Q1 | USA = 7, Romania = 4, Canada = 1, Israel = 1, Greece = 1, Ukraine = 1, France = 1, Chile = 1, Japan = 1, Democratic Republic of the Congo = 1 | 1993 - 2012 | Children aged 0-17 years living in alternative care for a minimum of 2 months (children's homes and foster care) |
| Boyle, 2015 | Systematic Review | 11 (4) | Q1, Qa | UK = 11 | 2002 - 2013 | Children in non-kinship permanent foster care, with formally organized contact mediated by fostering services |
| Eastman, 2019 | Systematic Review | 18(18) | Q1 & Qa | n.r | 2011-2017 | Young parents with CPS involvement |
| Wright, 2019 | Systematic Review | 38 (38) | Q1 | Europe = 14, Africa = 7, Middle East = 4, North America = 4, Asia = 3, Central America = 3, Global = 2, South America = 1 | 1972 - 2018 | Youth who resided in institutional care between the ages of 0â€“18 years, and resided full-time within institutional care. |
| Gillum, 2016 | Literature Review | 24 (24) | n.r | n.r | n.r | Individuals who experienced foster care and were college students or college graduates |
| Osei, 2016 | Rapid Review & Meta-analysis | 13(13); Review studies (5) | Q1 | USA = 13 | 1990 - 2013 | Youths ages 10-18 |
| Stewart, 2013 | Literature Evaluation | 27 (27) | Q1 | USA = 17, UK = 3, Sweden = 3, Australia = 1, Canada = 1, Germany = 1, Norway = 1. | 2001 - 2011 | Children and adolescents involved with child welfare services |
| Lutman, 2016 | Rapid Review | 22 (22) | Qa | n.r (but search was limited to UK, Ireland, USA, Canada, New Zealand, Denmark, Norway, Sweden) | 1995-2014 | Children (aged up to 18) living in foster family care (including kinship care) |
| Häggman-Laitila, 2019 | Systematic Review | 13(13) | Qa | USA = 9, Israel =1, India =1 | 2010-2017 | Young adults or adults who had been in foster care and had left the care |
| Lloyd, 2018 | Systematic Review | 7 (7) | Q1 | n.r | 2003 - 2012 | Mothers with substance use disorders whose children are is foster care |
| Steels, 2017 | Critical Review | 7(7) | Q1, Qa | Australia = 2, UK = 1, Ghana = 1, Romania = 1, Spain = 1, Sweden = 1 | 2008 - 2015 | Children and workers in residential care homes |
| Rock, 2015 | Systematic Review | 58 (58) | Q1, Qa | UK = 22, USA = 21, Canada = 7, Australia = 5, The Netherlands = 2, Sweden = 1. | 1960 - 2009 | Children in foster-care |
| DeLuca, 2019 | Meta-analysis | 25(6) | Q1 | USA = 7, Canada = 1, UK = 1 Australia =1, Spain =1, n.r =1 | 1983 - 2012 | Adolescents in foster care |
| Brown, 2014 | Literature Review | n.r | n.r | n.r | n.r | Looked after children who are no longer able to live with their parents and reside with extended family members and friends in kin placements compared to looked after children in non-kin care whose placements are provided or purchased directly by the local authority |
| Chodura, 2021 | Systematic Review & Meta-analysis | 43(43) | Md | USA =29, Other = 14 | 1994-2019 | Children in foster care |
| Milde, 2021 | Systematic Review | 6 (3) | Q1 | Sweden = 5, Finland = 1 | 2002-2018 | Former and current child welfare services clients (aged 2-18 years) in Nordic countries |
| Vanderwill, 2021 | Systematic Review | 29(29) | Q1, Qa, Md | USA = 29 | 2003 - 2017 | Children and youth in care. |
| Poon, 2021 | Meta-analysis | 9(9) | Q1 | USA = 9 | 2010-2019 | Youth currently and/or formerly engaged in the foster care system in the USA. |
| Saarnik, 2021 | Systematic Review | 24 (24) | Q1, Qa, Md | USA = 6, Canada = 4, Sweden = 4, UK = 3, Australia = 3, Spain = 2, Belgium = 1, Germany = 1 | 2000 - 2019 | Foster parents or children |
| Poitras, 2022 | Systematic Review | 18(18) | Q1 | USA = 7, UK = 4, Australia = 2, Canada = 2, Israel = 1, The Netherlands = 1, and Norway = 1 | 1985 - 2018 | Children aged 0 to 18 years placed in a foster family or in kinship family. |
| Nuñez, 2022 | Systematic Review | 12(12) | Q1 | USA = 12 | 2009-2019 | Foster youth, including foster alumni |
| Hassall, 2021 | Systematic Review & Meta-analysis | 31(31) | Q1, Qa | USA, The Netherlands, Sweden, Korea, Israel, Ireland, Australia (Numbers not reported) | 1991-2017 | Children aged 0-18 living in kinship care |
| Seker, 2022 | Systematic Review & Meta-analysis | 19 (19) | Q1 | Europe = 10, USA = 8, Oceania = 1 | 1996 - 2019 | Adults with a history of foster care or residential care in childhood or adolescence. |

*Number of studies relevant to our objectives; n.r = not reported; Q1 = quantitative; Qa = qualitative; Md = mixed design
