## Supplementary material for "Mapping the Evidence on Outcomes of Childhood Out-of-Home Care: A Scoping Review of Reviews": S1 File

**Factors associated with childhood out-of-home care entry and re-entry in high income countries: A systematic review of reviews**

1. **Search strategy**

A comprehensive search to identify peer-reviewed review articles was performed in EBSCOhost (covering MEDLINE, Education Research Complete, APA PsycArticles, APA PsycInfo, and CINAHL Ultimate), ProQuest (including Applied Social Sciences Index & Abstracts (ASSIA), Criminal Justice Database, Education Database, and Social Science Database), Cochrane Database of Systematic Reviews, and Epistemonikos. Grey literature databases were not searched as they typically contain primary literature, whereas this study focuses exclusively on peer-reviewed evidence reviews. The search strategy was initially developed by R.O. and was refined by J.N., K.H., S.B., as well as contributions from PPI members. Additionally, the expertise of a health librarian was leveraged to enhance the strategy.

The strategy was designed to be intentionally broad, incorporating terms related to or describing child social care, thereby minimising the risk of omitting relevant reviews. Also, additional search terms suggested by care experienced children and young people as part of public involvement expanded the scope of the literature search to include terms such as "residential care" and “care leavers.” The detailed search strategy employed for EBSCOhost databases is available in Table A1 (see appendix). Review filters available within each database were used to refine our results. In cases where pre-specified review filters were not available, searches included terms specifically related to review articles to ensure comprehensive coverage. This approach aimed to ensure that the search was both exhaustive and precise, facilitating the identification of relevant literature for inclusion in the review.

**2. Study selection**

Covidence software was used to organise and screen the identified records, ensuring the automatic and manual removal of duplicate entries. Six reviewers (R.O., N.J., M.J., M.P., N.K., and K.C.) independently screened the records. Each record was assessed by two reviewers, who categorised it as 'yes,' 'no,' or 'maybe.' Two supervising reviewers (S.B and K.H) resolved conflicts and ‘maybe’ responses. Reasons for exclusion were documented at this stage, and records receiving a ‘yes’ vote proceeded to full-text screening. In the subsequent phase, two reviewers independently assessed the full texts of each record, classifying them as ‘include,’ ‘exclude,’ or ‘maybe.’ The same conflict resolution strategy was applied during this phase. A record of all decisions was maintained, including the reason for each exclusion. Finally, references for all included articles were exported to Mendeley reference manager. The search results and study selection process were depicted using the PRISMA flow diagram (Figure 2) (Moher et al., 2015).

**3. Patient and Public Involvement**

The review of reviews was conducted with input from the Welsh National Centre for Population Health and Wellbeing Research PPI group through grant development, the review and interpretation of findings. This ensured the questions, design and interpretation of findings were relevant to and reflective of the lived experiences of people involved with the child welfare system. The public involvement process engaged two key groups: children and young people with care experience (15-25 year-olds) through CASCADE Voices, a research advisory group for care experienced young people, and parents who have had a child removed from the home.

**Appendix**

**Table A1. Search Strategy in EBSCOhost Databases (MEDLINE, Education Research Complete, APA PsycArticles, APA PsycInfo, CINAHL Ultimate)**

| Step | Query | Limiters/Expanders | Results |
| --- | --- | --- | --- |
| 1 | "foster care" OR "foster home" OR "foster family" OR "foster parent" OR "foster carer" OR "substitute family" OR "family foster home" OR "kin* care" OR "child* in care" OR "out#of#home care" OR "looked#after" OR "child* in need" OR "vulnerable child*" OR "social service*" OR "child welfare" OR "residential care" OR "group home*" OR "relative  care" OR "guardian care" OR "care experience" OR "care leaver" | Limiters - Publication Date: 2013/01/01-2024/01/31 | 129,301 |
| 2 | TI "foster care" OR "foster home" OR "foster family" OR "foster parent" OR "foster carer" OR "substitute family" OR "family foster home" OR "kin* care" OR "child* in care" OR "out#of#home care" OR "looked#after" OR "child* in need" OR "vulnerable child*" OR "social service*" OR "child welfare" OR "residential care" OR "group home*" OR "relative  care" OR "guardian care" OR "care experience" OR "care leaver" | Limiters - Publication Date: 2013/01/01-2024/01/31 | 127,548 |
| 3 | TI "foster care" OR "foster home" OR "foster family" OR "foster parent" OR "foster carer" OR "substitute family" OR "family foster home" OR "kin* care" OR "child* in care" OR "out#of#home care" OR "looked#after" OR "child* in need" OR "vulnerable child*" OR "social service*" OR "child welfare" OR "residential care" OR "group home*" OR "relative  care" OR "guardian care" OR "care experience" OR "care leaver" | Limiters - Full Text; Peer Reviewed; Publication Date: 2013/01/01-2024/01/25  Narrow by Methodology: -metasynthesis -meta analysis -systematic review -literature review  Narrow by Language: -English  Narrow by Subject Major: -foster care -social support -foster home care -social services -family -residential care -child welfare | 213 |
